# Breast cancer over-diagnosis due to mammography screening – A long-term follow-up population study of BreastScreen Norway

**DOI:** 10.64898/2026.06.02.26354696

**Authors:** Torunn Heggland, Lars Johan Vatten, Signe Opdahl, Harald Weedon-Fekjær

## Abstract

**Objectives:** Estimates of breast cancer over-diagnosis related to mammography screening varies substantially. Over-diagnosis is commonly defined as cases that would not have been detected during the person’s remaining lifetime in the absence of screening. We here aim to quantify over-diagnosis in the population-based BreastScreen Norway mammography screening program using long-term follow-up and more detailed modeling than previous studies.

**Setting:** We applied data on Norwegian screening patterns and breast carcinoma incidence for the period 1987-2019, covering women aged 49-84 years, leveraging the gradual implementation of the organized biennial BreastScreen Norway screening program for women aged 50-69 during 1995-2005.

**Methods:** Using an extended age-period-cohort model, we estimated excess lifetime risk of invasive breast cancer and ductal carcinoma in situ in the presence of program screening, as an indicator of over-diagnosis among screen-detected cases.

**Results:** Lifetime risk of breast carcinomas was 6.6% (95% confidence interval 2.5% to 10.7%) higher for invited than for non-invited women. This indicates that 18% (95% confidence interval 7.3% to 28.0%) of screen-detected cases may be over-diagnosed, and that approximately one in 86 (95% confidence interval 54 to 210) screened women were over-diagnosed during their screening period. Using effect estimates from previous studies, we estimated that approximately three women are over-diagnosed for every breast cancer death prevented by screening, and that 87% of over-diagnosed tumors might grow extremely slowly.

**Conclusions:** Over-diagnosis related to mammography screening is a considerable problem, but its extent may be smaller than reported in some previous studies. Most over-diagnosed tumors likely grow very slowly.

## Introduction

The aim of mammography screening is to detect breast cancer in its preclinical stage and thereby improve prognosis due to more effective treatment. Earlier detection does, however, also entail a risk of over-diagnosis, defined as breast cancer that would not have been detected within the woman’s remaining lifetime in the absence of screening ^1^. Because most women diagnosed with breast cancer at screening receive treatment, it is not possible to know if the tumor would have become clinically detectable later, and in consequence the level of over-diagnosis cannot be directly observed. Estimates of over-diagnosis range from zero to more than 50% ^2–7^. Some of that variation is due to different measures of over-diagnosis ^8^, but there is also a large variation due to different analytical strategies and assumptions ^9^. The ideal analytical approach would be to compare long-term breast cancer incidence in the presence and absence of screening, preferably in a randomized screening trial. However, in all randomized trials to date, most women in the control group were offered screening after some time, precluding long-term follow-up of sufficiently large unscreened populations ^10^. Due to the lack of suitable trial data, an alternative approach is to follow the introduction of an organized screening program and compare breast cancer incidence between women in regions that were first invited and women in regions that were not yet invited.

In Norway, the national mammography screening program targeting women 50-69 years of age (BreastScreen Norway) was gradually introduced over a period of almost 10 years. By carefully comparing breast cancer incidence between counties where women had been invited, and counties where women had not yet been invited, most of the incidence trend differences are likely to reflect screening effects. We took advantage of this gradual introduction of screening, combined with long-term follow-up and detailed statistical modeling, to estimate the extent of over-diagnosis in BreastScreen Norway.

## Methods

### Study population and data sources

The study included all women younger than 85 years of age living in Norway during the period 1987-2019. The women contributed person-years until they were censored due to death or end of follow-up. For each of the 18 Norwegian counties, we used annual data on population size and first diagnosis of invasive breast carcinomas or ductal carcinoma in situ (DCIS), collectively referred to as breast carcinomas, aggregated by one-year age groups from the Cancer Registry of Norway. In the analysis, the women’s age was deduced from the birth cohort and calendar year. It is mandatory to report all findings of breast carcinoma to the Cancer Registry of Norway, and only 0.2% of all breast cancers are ascertained from death certificates alone ^11^. Reporting of DCIS became mandatory from 1993, but the stable DCIS incidence rates around 1993 suggest that registration was almost complete even before mandatory registration was introduced (Supplement Table 1). We applied all-cause mortality among women in Norway by age for 2019 from Statistics Norway.

BreastScreen Norway invites women aged 50-69 to mammography screening every two years ^12^. The Cancer Registry administers the program in which women, based on their respective birth cohort, are invited to county-wide screening rounds that last for approximately two years. In the analysis, we used the exact starting and ending dates of invitations of the birth cohorts. The program started in late 1995 as a pilot involving four large counties, comprising approximately 40% of eligible women in Norway. Women in the entire age span were invited from start of the program. Between 1999 and 2005 the program was gradually expanded to include the remaining 14 counties. The overall attendance to the program has been relatively stable at around 76% ^12^. When the program started, screen-film mammography was used, and full-field digital mammography was gradually introduced from 2000 ^13^.

In the modeling, we wanted to correct for factors that may have caused a rapid change in the incidence of breast cancer. At the same time as the introduction of BreastScreen Norway, there was extensive use of menopausal hormone therapy ^14^, which is known to increase the risk of breast cancer ^15^. However, after the initial results of the Women’s Health Initiative (WHI) trial were published, suggesting that hormone therapy might also increase the risk of cardiovascular disease ^16^, the use decreased sharply from around 2002 (Supplement Figure 1). To avoid any bias due to changes in hormone therapy consumption, we applied county-specific sales figures of preparations containing estrogen (G03C according to the Anatomical Therapeutic Chemical classification ^17^) and estrogen-progestogen combinations (G03F) from the Norwegian Drug Wholesales Statistics. We scaled the sales figures according to the age distribution among prescription users of the respective hormone therapies registered in the Norwegian Prescribed Drug Registry. Both sources of menopausal hormone use are available from the Norwegian Institute of Public Health (See Supplement Material p 1).

### Statistical analysis

We used Poisson regression analysis to estimate the effects of organized screening on breast carcinoma incidence, exploiting the contrasts in breast carcinoma incidence between counties that initiated screening at different time periods. When applying Poisson regression, aggregating individual data based on the least common variable combination of the analysis does not alter the final estimated values. Data were therefore first aggregated internally in the Cancer Registry of Norway to address practical and data privacy issues.

In the basic breast carcinoma incidence model, we included age, period, and cohort variables, as age-period-cohort (APC) Poisson regression models are known to efficiently capture variations in cancer incidence ^18, 19^. As incidence changes typically appear gradually ^20^, we used natural cubic splines to smooth the effects of age (inner knots at age: 50, 52, 54, 56, 60, 70, and 80), period (inner knots at year: 1997 and 2009) and birth cohort (inner knots at birth cohort: 1929 and 1943), to limit the number of variables in the model. For the age variable we chose more frequent knots early in the age span to increase flexibility around the age of menopause, whereas the knots for period and birth cohort were evenly distributed over the respective spans. We included a variable for hormone therapy use given by mean daily doses with a one-year time lag ^14^, to reflect a reasonable time lag between use of hormone therapy and increased risk of breast carcinoma ^15^. We also allowed for different breast carcinoma incidence levels between counties.

Similar to our previous modelling work ^14, 21, 22^, we extended the model with separate variables to take into account that successive rounds of screening may have different effects. Thus, the first screening of a given birth cohort is likely to increase the incidence more than subsequent screening rounds, as no breast carcinomas have previously been detected by organized screening in the given birth cohort. The subsequent screening rounds will primarily detect newly developed preclinical cases, thereby shifting breast cancer detection to younger ages. However, we handled the second screening round separately to account for possible first-time participation and cases missed in the first round. We therefore included indicator variables to cover the first screening round around age 50-51, the second screening round around age 52-53, and the subsequent screening rounds from approximately 54 years. For screening rounds starting or ending during the calendar year, these variables constituted the proportion of the calendar year women in a birth cohort were invited to the respective screening rounds. Such a use of proportions in Poisson regression only induces a negligible bias ^23^. We also applied natural cubic splines to allow for non-linear effects by age for the screening period (inner knot at age: 60). When screening ceases, the breast carcinoma incidence is likely to decrease because many of the cases that would have become clinical at that age were already detected in the screening program. We included a variable for no longer being covered by the program, and allowed for a gradually declining effect of screening, using natural cubic splines for the time since screening cessation.

As we aimed to model breast carcinoma incidence in the fully implemented program, we excluded data on the screening initiation at higher ages. In practice, data reflecting combinations of calendar year, birth cohort and county that corresponded to a first screening invitation above 53 years of age, or a second screening invitation for women above 55 years of age, were not used in the modelling.

Since some breast cancer risk factors differ between younger and older women ^24^, we fitted a separate model for the pre-screening ages (0-48 years), which was used in the calculation of lifetime risk. The full specifications of both incidence models are given in the appendix (Supplement Material p 2). To avoid potential unconscious influence from personal beliefs regarding over-diagnosis level, we also specified a pre-hoc incidence model before running any analysis (Supplement Material p 4).

#### Calculation of over-diagnosis

Using our incidence model combined with all-cause mortality among Norwegian women, we estimated over-diagnosis as the added breast cancer lifetime risk in the presence of organized screening. We calculated overdiagnosis as a proportion of screening-detected cases, and relative to all cases in the absence of screening, across different age ranges (0-84, 50-84 and 50-69 years).

In the estimation, we expressed the quantities as breast carcinoma risks (Fig 1). From the incidence model, we estimated age-specific risk of a first breast carcinoma among all women alive at the given age, both in the presence and absence of screening. Further, we calculated the lifetime risk of breast carcinoma (i.e., 0-84 years), and the total risk of breast carcinoma across different age ranges (i.e., 50-84 and 50-69 years), by combining the age-specific risks and the probability of being alive (Supplement Material p 4). The numerator in our over-diagnosis measures was thus the difference in lifetime risk between women invited, and women who were not invited, to screening. For the estimation of screening-detected cases, we applied the observed proportion of screening-detected cases among breast carcinomas in women aged 50-69 in 2016-2019 (68%), and multiplied that proportion with the breast carcinoma risk across ages 50-69 years in the presence of screening. To enable comparison with studies with shorter follow-up time, we also used the incidence model to estimate over-diagnosis with shorter follow-up time.

**Figure 1:**
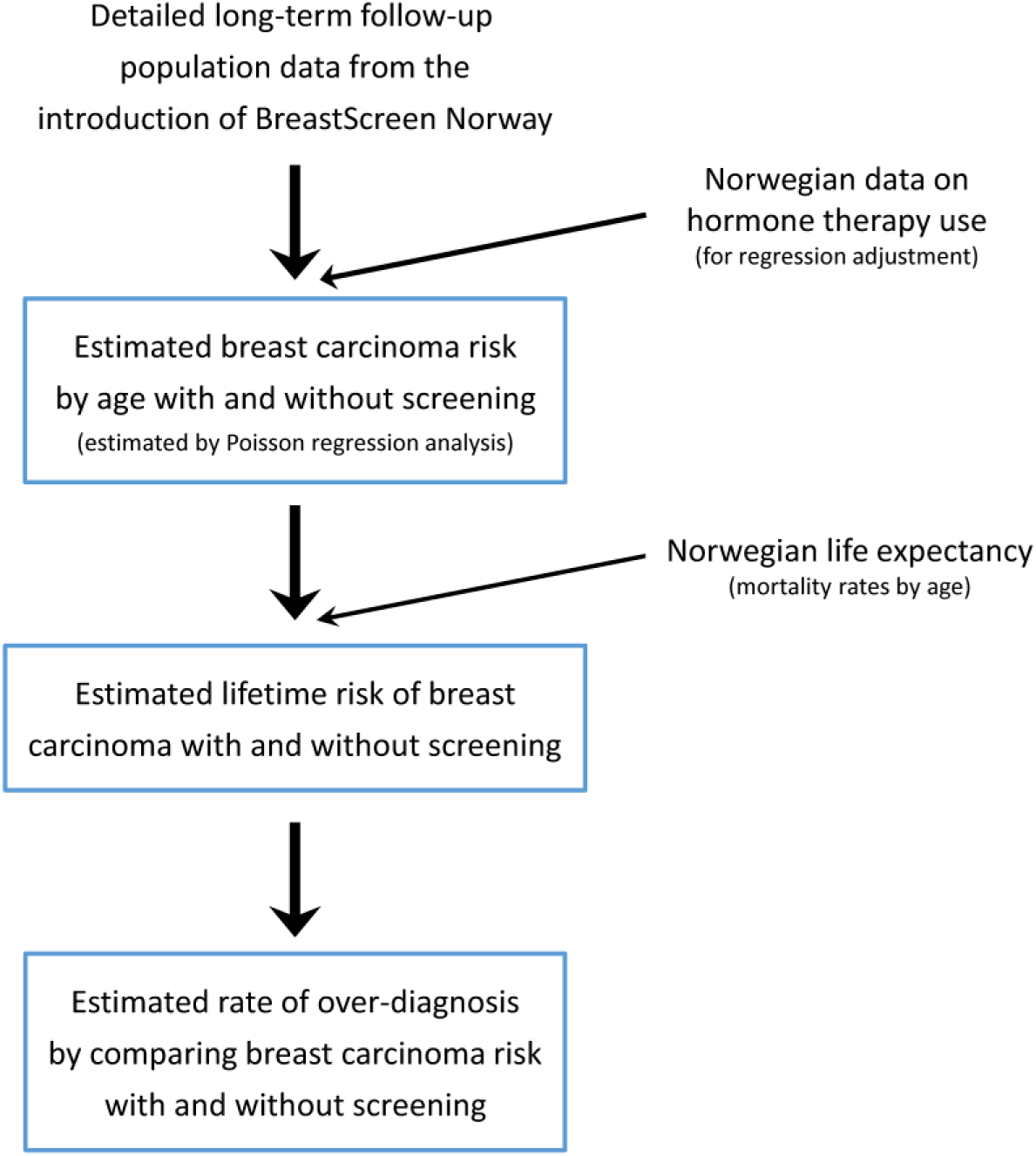
Outline of the method used for over-diagnosis estimation and applied data.

For over-diagnosis calculations we applied the incidence model and all-cause mortality on the 1969 birth cohort for calendar year 2019, with hormone therapy set at the national level of 2019. The over-diagnosis estimates are weighted averages across all Norwegian counties.

For sensitivity analyses, we either removed the hormone variable, the period variable or the interaction between age and subsequent screening. As even 15 years of observation past the last screening exam might not cover all potential lead time, we added a sensitivity analysis extending the observed compensatory drop in breast cancer incidence up to 100 years of age, using two approaches: i) We extended the observed difference at 84 years of age in breast cancer incidence with and without screening, and ii) We extended the observed difference at 84 years (as in i) combined with extending the observed slope during the age range 80-84 years. To enable comparison with the main analysis, we kept the lifetime risk up to 84 years of age as the denominator in the sensitivity analysis.

Akaike’s information criteria (AIC) was used in the selection of the appropriate incidence model.

To assess statistical uncertainty, we calculated 95% bootstrap percentile confidence intervals (CI) based on 10 000 repetitions. All statistical analyses were conducted using the R statistical package (version 4.2.2, R Foundation for Statistical Computing, Vienna, Austria) ^25^.

## Results

The analysis included 63 378 cases of invasive breast carcinomas or DCIS among 23 709 444 person-years of observation, in the age range 49-84 years. Among these, 3096 cases occurred in 2019 during 951 702 person-years of follow-up. We estimated a lifetime risk of breast carcinoma of 14.4% among women invited to screening, and 13.5% in the absence of screening.

Overall model fit of the APC model was good, with a small over-dispersion (1.03). In the absence of screening, the fitted APC model showed that breast cancer incidence increased gradually by age with the expected plateau around menopause ^14^ (Fig 2 and Supplement Material p 3). With screening present, there was a marked peak in breast cancer incidence around the initial screening, an elevated incidence with continued screening, and a substantial reduction when invitations to screening ceased. Thereafter, it gradually approached the incidence in the absence of screening.

**Figure 2:**
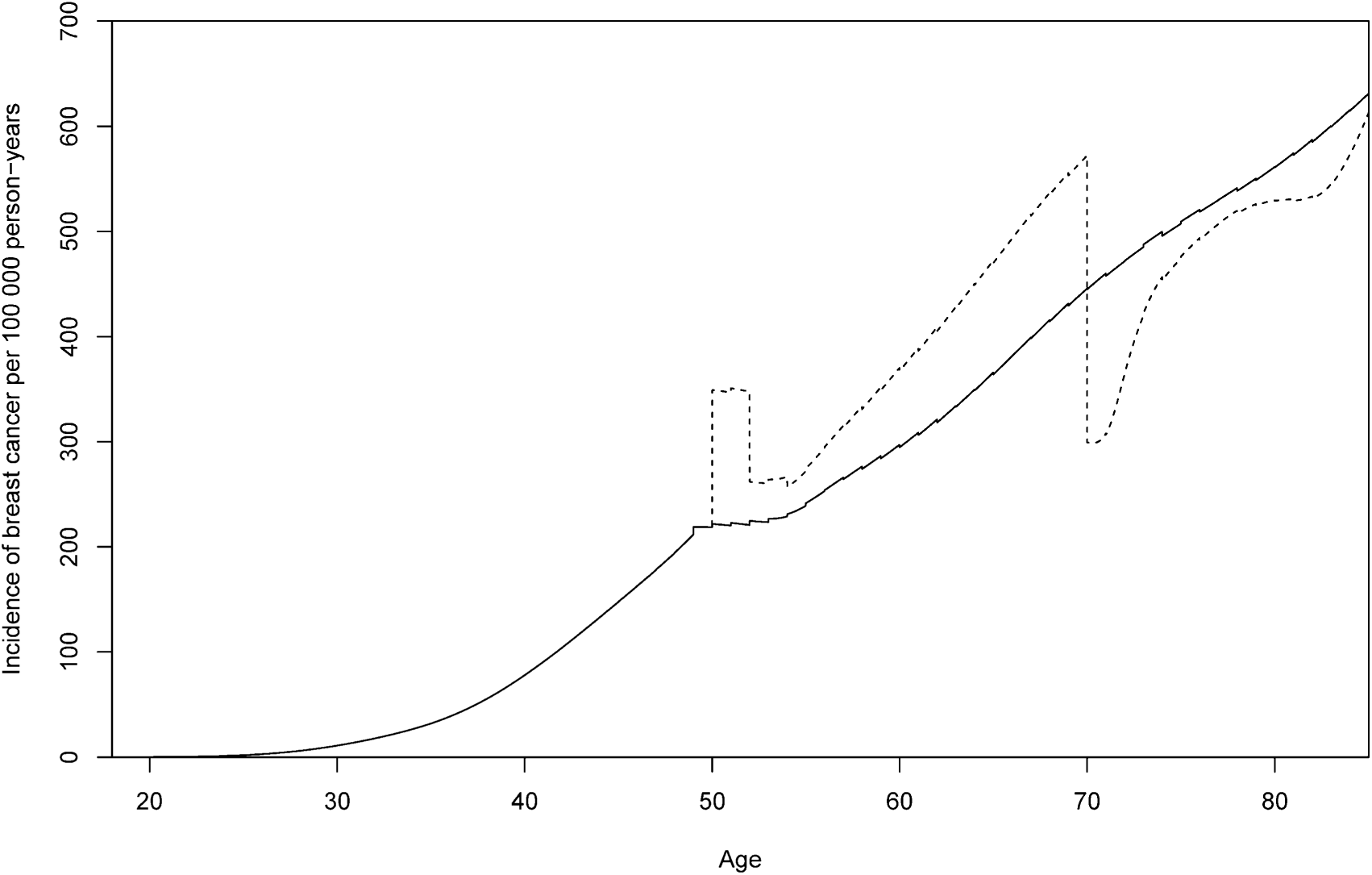
Estimated incidence rate of breast cancer by age in absence (solid line) and in presence (dotted line) of invitation to BreastScreen Norway (estimated for year 2019 with average hormone therapy use). The rectangular shape at age 50 years in the presence of screening arises as women are invited gradually over a two year period, spreading out the initial screening effect.

We estimated that 18.0% (95% confidence interval 7.3% to 28.0%) of screening-detected breast carcinomas were over-diagnosed. The pre-hoc model specified before running any analysis, gave an almost identical over-diagnosis estimate among screening-detected cases of 18.3% (95% confidence interval 7.7% to 28.1%). Women invited to screening from age 50 to 69 years had a lifetime risk of breast cancer (up to 84 years of age) that was 6.6% (95% confidence interval 2.5% to 10.7%) higher than that of women not invited (Table 1).

**Table 1:**
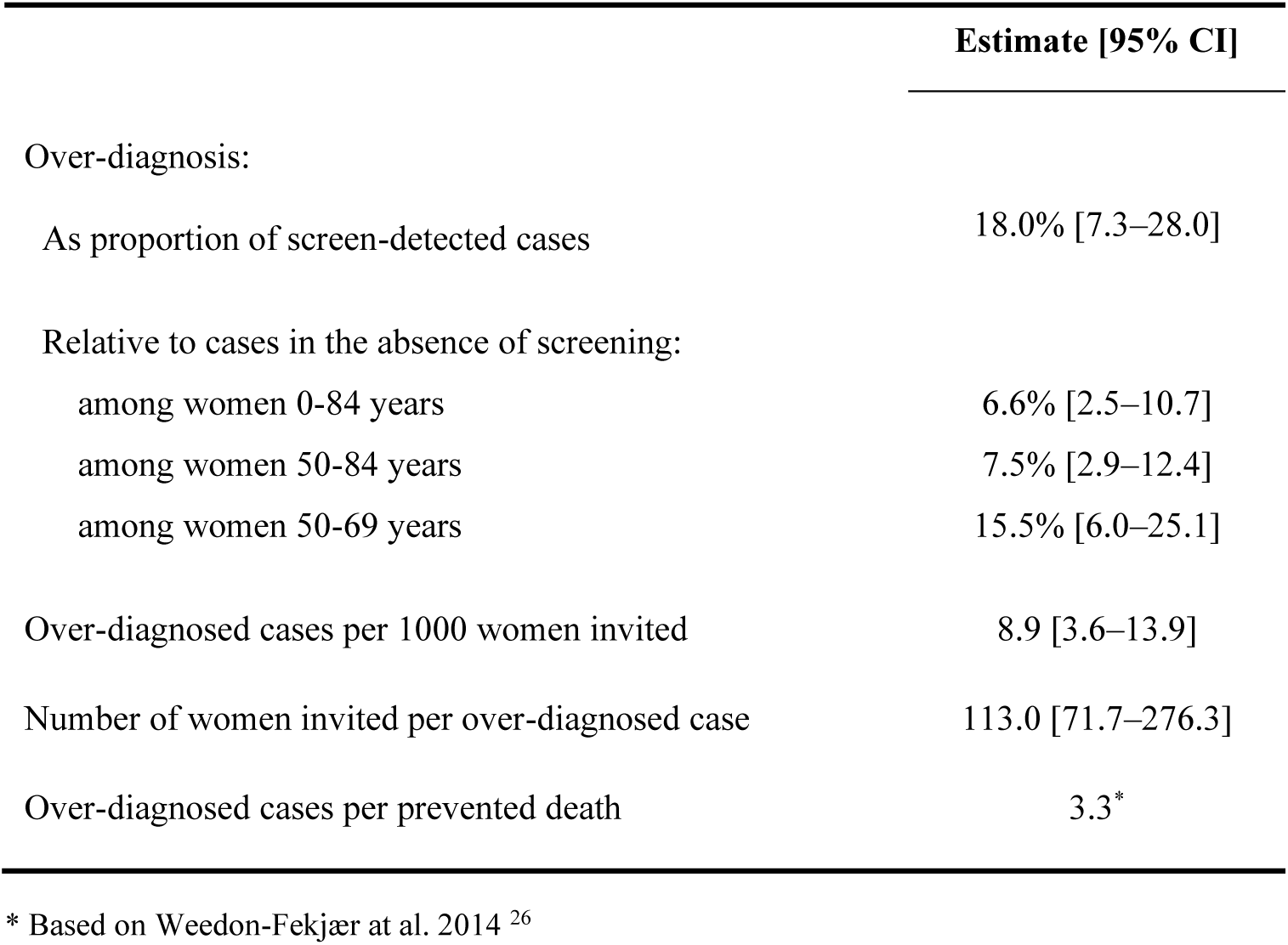
Estimated over-diagnosis after implementation of BreastScreen Norway.

We estimated the rate of over-diagnosis to 9 cases (95% confidence interval 4 to 14) per 1000 women invited to the full program, corresponding to 113 (95% confidence interval 72 to 276) women invited per over-diagnosed case. Assuming 76% attendance, approximately one woman will be over-diagnosed among every 86 (95% confidence interval 54 to 210) women attending the screening. Combining this estimate with results from our earlier study of breast cancer mortality ^26^, we estimated that approximately three women would be over-diagnosed for each death prevented by screening (Table 1). With similar methodology as in the present work, we previously estimated that 15.7% of screening-detected cases in the BreastScreen Norway program would not have become clinical by 85 years of age (if the women had lived to that age) ^22^. Using this previous result, we estimated that 87% (15.7/18) of over-diagnosed cases were due to extremely slow-growing carcinomas that would not have become clinical within the first 15 years following cessation of the screening program.

Sensitivity analyses showed some variations with different model specifications; however, the applied incidence model gave the best fit according to AIC (Table 2). Limiting the follow-up after screening cessation, we found the estimates of over-diagnosis to be somewhat higher (Table 3). Extending the duration of the observed compensatory drop in breast carcinoma incidence beyond 84 years of age had little impact on our estimated overdiagnosis rate, even when including lifetime risk up to 100 years of age. Incorporating both the difference at 84 years of age and the 80-84 years slope, the increase in lifetime risk of breast carcinoma under screening dropped from 6.6% to 5.7% (using the same denominator, i.e., lifetime risk up to 84 years of age, for easier comparison). Incorporating only the difference at 84 years of age, the increase in lifetime risk under screening decreased to 5.5%.

**Table 2:** Over-diagnosis estimates and AIC values from different APC-models.

| Model | Over-diagnosis as proportion of screen-detected cases | Over-diagnosis relative to cases in the absence of screening, among women aged 0-84 |
| --- | --- | --- |
| Main model† | 18.0% | 6.6% |
| Pre-hoc model‡ | 18.3% | 6.7% |
| Sensitivity analysis: (modifications of main model) |  |  |
| Without age trend for subsequent screening | 12.8% | 4.5% |
| without hormone therapy variable | 17.7% | 6.6% |
| without period variable | 17.3% | 6.3% |
| Only invasive cancers\|\| | 8.4% | 2.6% |

\* Lower AIC indicates a better model fit.
† Specifications of natural cubic splines knots for main model: Age: Knots at 50, 52, 54, 56, 60, 70 and 80 years of age. Period and cohort: Three degrees of freedom with knots at quantiles. Time since screening: Knots at 1, 2, 5, 10 and 13 years. Subsequent screening rounds: Boundary knots at ages 53 and 72 years, and inner at 60 years of age.
‡ The main model with linear effect for subsequent screening by age, no inner knot at 1 year for time since screening, and data for women 48-84 years of age.
§ AIC for the pre-hoc model (AIC=74978) and the analysis with only invasive cancer (AIC=71488) are not comparable due to different input data.
|| The invasive only estimate, calculated ignoring all DCIS cases, should be interpreted with care as DCIS detection at screening might hide later invasive cancers.

**Table 3:**
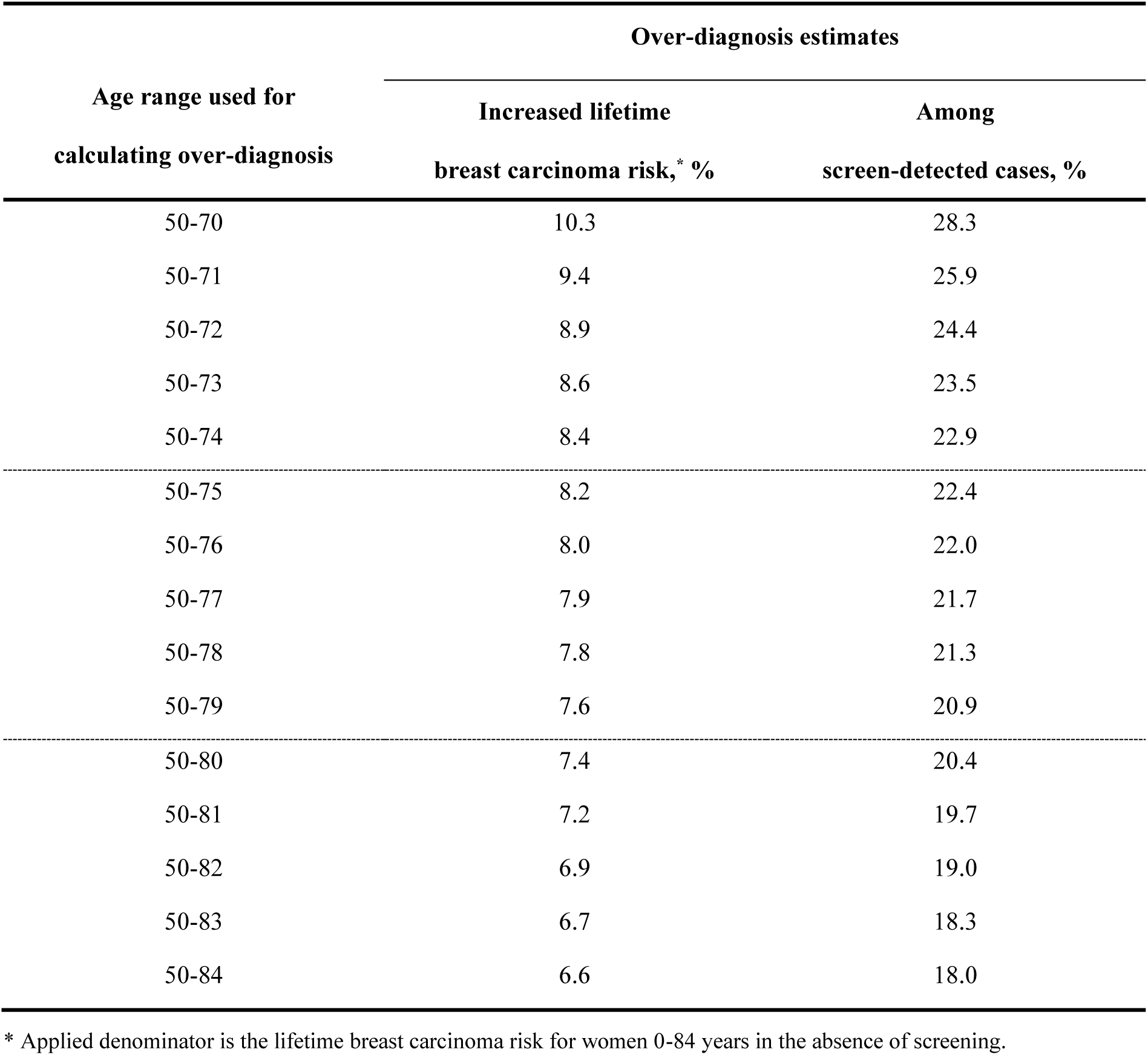
Effects of limited follow-up after screening cessation; over-diagnosis estimates with artificially shorter post screening follow-up.

## Discussion

With long-term follow-up and detailed modelling, we estimated that 18% of screening-detected breast cancers were over-diagnosed in this public biennial screening program for women aged 50-69 years. We also found that approximately three women were over-diagnosed for each death prevented by screening.

To avoid bias due to selective attendance, the analysis was based on invitation to screening and not on screening attendance. There are methods to adjust for self-selection bias ^27, 28^; however, the risk differences between attenders and non-attenders may have changed over time, due to variations in factors such as ethnicity composition in the population and opportunities for non-organized screening. Over-diagnosis, and excess incidence with screening present, is however only related to the attending women. Hence, an estimate of over-diagnosis associated with invitation to screening can easily be converted to exclusively cover attending women.

We fully utilized all detailed information on timing of screening invitations in Norway to compare breast cancer incidence between invited and not yet invited women. Although the gradual inclusion of counties was not truly randomized, we could follow the stepwise implementation of screening that happened at different time points. Thus, we could separate the likely effect of screening from that of underlying fluctuations in breast cancer incidence. Our model makes no assumptions about the direction and strength of the different screening effects, yet the estimated screening effects fit very well with presumed effects of screening (Fig 2).

Detection of preclinical breast carcinomas by screening induces lead-time; meaning the time interval between screening detection and the time when clinical detection would occur in the absence of screening. Taking proper care of lead-time is a challenge in over-diagnosis studies, because it requires either unverifiable assumptions about lead-time or unusually long follow-up. However, in this long-term follow-up study we could observe women until 84 years of age, probably taking most lead-time^29^ into account, as indicated by the moderate change in the overdiagnosis estimate when extending the observed slope up to 100 years of age. The effect of extending follow-up past 84 years of age is limited due to very few women remaining alive in the higher age ranges.

We estimated over-diagnosis for DCIS and invasive breast cancer combined. DCIS is a precancerous lesion that is often asymptomatic, and its detection is therefore closely linked to screening. After the introduction of screening, DCIS accounted for approximately 20% of screening-detected cases ^31^. Because DCIS may develop into invasive disease, its treatment is similar to that of early stage invasive breast carcinoma. As treatment of DCIS may to some extent prevent later invasive disease, we cannot reliably distinguish DCIS-related over-diagnosis from that of invasive breast carcinoma.

Our estimate indicates that most over-diagnosed cases in BreastScreen Norway are likely to be extremely slow-growing tumors (87%). This estimate is influenced by the high Norwegian life expectancy (84.7 years for women in 2019), and a moderate upper screening age of 70 years. Hence the results might not be directly generalizable to populations with shorter life expectancy or screening continued into higher age ranges.

It is difficult to know exactly how the hormone therapy “wave” in the 1990s affected breast cancer incidence and the level of over-diagnosis. Use of menopausal hormone therapy increases mammographic density and may therefore have a masking effect on mammography ^32^. Also, users of hormone therapy may be more likely to participate in screening than non-users ^33^. However, to limit the number of free model parameters, we did not include all these possibilities in the analysis using one overall hormone therapy variable.

It seems likely that the sensitivity of the screening program has increased over time, both due to increased reader experience, and due to the transition from analogue to digital mammograms. In their 2020 review, Farber et al. ^34^ found that digital mammography detects 10% more breast carcinomas, with an especially large increase in DCIS. In our study design, a higher sensitivity would immediately affect ongoing screening, while an increased compensatory drop in incidence after cessation of screening would need more time to appear. As digital screening was introduced during our observation period, our data with a set observation period will be more affected by digital screening in the active screening phase than the post-screening phase. This might have resulted in a somewhat inflated level of over-diagnosis, as we for the digital screening cohorts might not have fully taken into account their likely larger compensatory drop post-screening. The use of digital breast tomosynthesis was limited during the study period.

A substantial proportion of women reported having had mammograms prior to their first BreastScreen Norway attendance ^35^, but the impact of these examinations is uncertain. There was, however, no clear increase in county-specific breast cancer incidence until the screening program was initiated in the respective counties (Supplementary File p 5). This indicates that the use of opportunistic mammography was reasonably constant pre-screening. Hence, the pre-programme opportunistic mammography might be seen as a background screening level prior to the public screening programme. In this sense, our estimates could be interpreted as overdiagnosis specifically due to the extra mammography exams following the introduction of the public programme.

### Comparison with other studies

Previous studies of excess incidence related to mammography screening typically reported rather high levels of over-diagnosis ^4, 5, 30, 36^. Jørgensen and colleagues estimated 33% excess breast cancer incidence in Denmark, but adjustment for incidence trends was less precise than in our study.^4^ Also using BreastScreen Norway data, Kalager and colleagues estimated 15-25% excess incidence of invasive breast cancer using different age groups and ad-hoc lead-time adjustments ^36^. Compared to our study, their follow-up was shorter, and the screening vs. non-screening groups were not precisely separated. Falk and colleagues compared participants and non-participants in BreastScreen Norway and estimated that 17-20% of all cases detected after 50 years of age may be over-diagnosed ^30^. However, studies that consider women’s attendance may be vulnerable to self-selection bias.

Ripping and colleagues applied an age-period-cohort model for breast cancer incidence in the Netherlands ^37^. They estimated that 14% of cases were over-diagnosed considering all breast cancer cases among women aged 20-99 years in the denominator. This is considerably higher than our estimate for comparable age groups. The study did, however, only include five years of observation after screening cessation, which may lead to an overestimated level of over-diagnosis. Njor and colleagues reported 2.3% excess cumulative breast cancer incidence in a Danish cohort aged 56-69 years at the start of biennial screening, where women were followed for at least 8 years after last screening invitation ^38^. Their estimate is lower than in our study, and might be biased downwards due to few screening rounds for a large proportion of the participants.

In the NHS Breast Screening Program in England, Blyuss and colleagues estimated 3.7% (95% confidence interval 0.8 to 17.4) over-diagnosis among screening-detected cases, using a case-control design ^39^. That estimate is considerably lower than in the present work, but the confidence intervals are overlapping. Over-diagnosis estimates tend to be rather low when estimated from lead-time models or simulation models ^3, 7, 40–42^. In the Netherlands, de Koning and colleagues estimated that 8% of screening-detected cancers may be over-diagnosed using the micro-simulation model MISCAN and data for 1989-2002 ^41^.

Michalopoulos and Duffy applied a combination of trend analysis and models for lead-time and estimated 15–17% over-diagnosis among screen-detected cases in BreastScreen Norway, which is more in line with our findings ^42^. In their lead time estimation approach, Michalopoulos and Duffy applied data from interval cancers to avoid bias from overdiagnosed cases. The strength of the lead time modelling approach is that it does not rely on a given maximum observable sojourn time. However, it depends heavily on the assumed exponentially distributed sojourn times. In some ways, our sensitivity calculations extending the compensatory drop up to 100 years of age are similar to the lead time modelling approach, as we also add some modelling assumptions to extend inference beyond the last observation. Reassuringly, the here given sensitivity analysis indicates that our estimates do not rely much on assumptions of what happens past our observation period.

Ryser and colleagues estimated 15% over-diagnosis among screening-detected cases in the US with biennial screening for ages 50-74 years, using a lead-time approach accommodating non-progressive cancers ^3^. Comparing these results with our findings, our excess-incidence study seems to be in closer agreement with lead-time and simulation approaches than most previous excess-incidence studies. Hence, this study aligns the results from the two different approaches, indicating that results of earlier excess incidence studies might be biased due to short follow-up and inadequate adjustment for underlying incidence trends.

In conclusion, using careful modelling of breast carcinoma incidence, detailed information about the introduction of screening, and long-term follow-up, we found that over-diagnosis is less prevalent than reported in many previous excess-incidence studies, but still corresponds to three women over-diagnosed for every breast cancer death prevented by screening.

## Statements and Declarations

### Ethical considerations

The applied input table for Poisson regression was calculated internally at the Cancer Registry of Norway. Hence, no ethical approval was needed for this study.

### Consent to participate

Not applicable

### Consent for publication

Not applicable

### Data availability

The data are available for research projects from the legal administrator of the data, the Cancer Registry of Norway. The interpretation and reporting of these data are the sole responsibility of the authors, and no endorsement by the Cancer Registry of Norway is intended nor should be inferred. For data requests use.

## Supplementary material

### Registration of DCIS

**Table S.1.** First cases of DCIS or invasive breast cancer relative to first cases of only invasive breast cancer, for all ages in Norway.

| Year | Incidence of first breast carcinoma (DCIS+invasive) relative to incidence of only first invasive breast carcinoma |
| --- | --- |
| 1987 | 1.034 |
| 1988 | 1.035 |
| 1989 | 1.035 |
| 1990 | 1.040 |
| 1991 | 1.039 |
| 1992 | 1.041 |
| New guidelines for DCIS registration |  |
| 1993 | 1.029 |
| 1994 | 1.037 |
| Screening initiation |  |
| 1995 | 1.042 |
| 1996 | 1.062 |
| 1997 | 1.072 |
| 1998 | 1.063 |

### Supplementary information regarding the hormone therapy variable

Electronically available national sales statistics for hormone therapy in the Norwegian Drug Wholesales Statistics dates back to 1987. County-specific sales statistics have been available electronically in four counties since 1992, and for all counties since 1999. For the years without county-specific sales statistics electronically available, we used statistics collected from paper lists for 1987, 1991, and 1995, combined with linear interpolation for the years in between.

The Prescription Database of Norway dates back to 2004. For the period 1987-2003, we used the age distribution for prescription users in 2004 as an approximation to the previous age distribution. A small proportion of hormone therapy was prescribed to men. This was taken into account when calculating the hormone therapy variable.

### Use of hormone therapy among women in the BreastScreen Norway target age range (50-69 years of age)

**Figure S.1.**
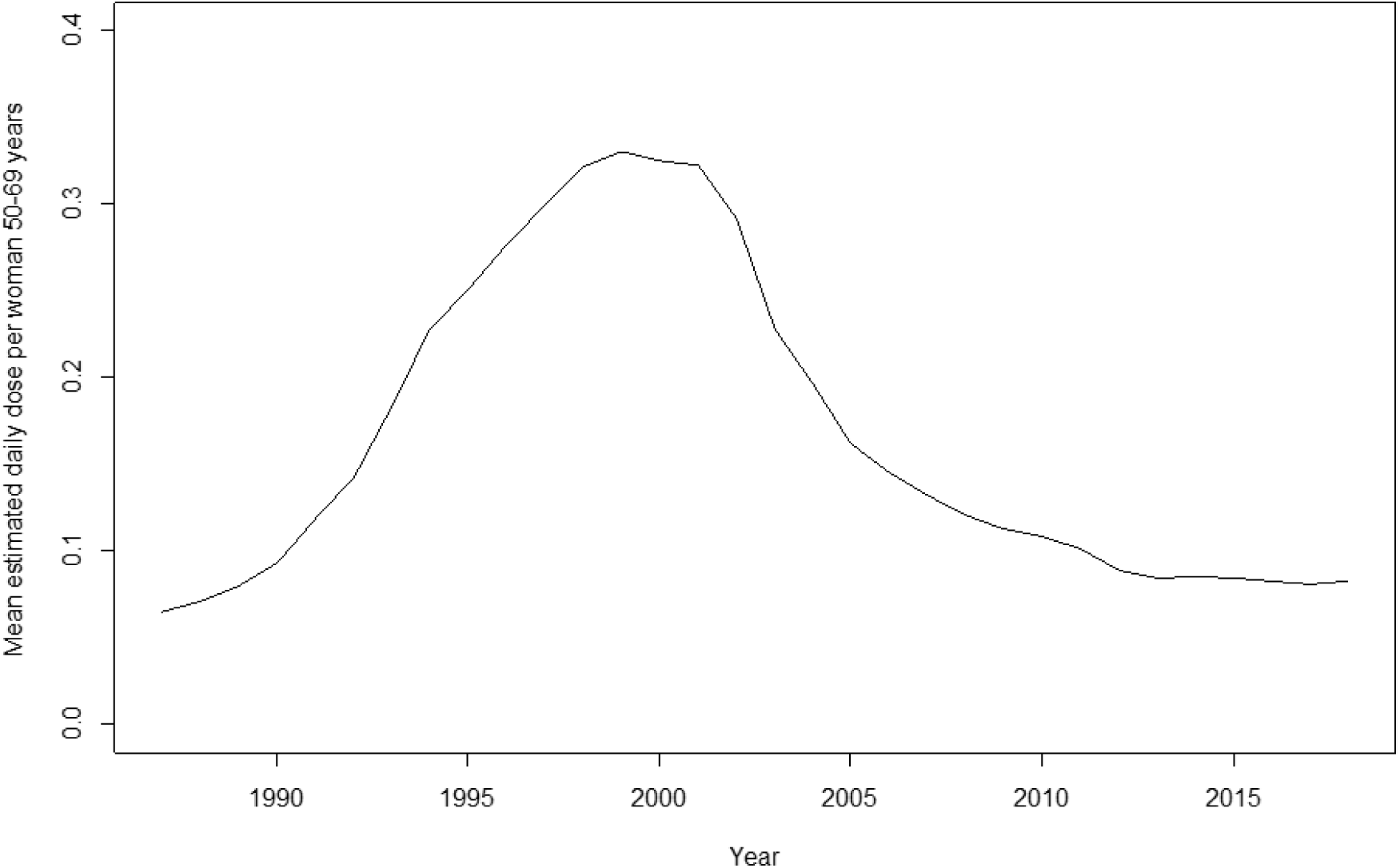
Mean daily dose of hormone therapy per Norwegian woman in the age group 50-69, calculated on the basis of total sales figures and age distribution among prescription hormone therapy users.

### The extended APC Poisson regression model for breast carcinoma incidence

The applied breast carcinoma incidence model from the age of 49 years could be written as

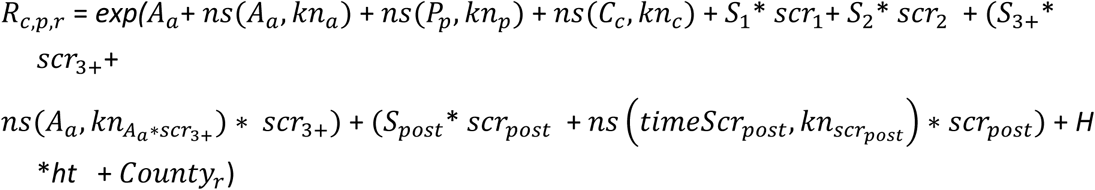

where *R*_*c*,*p*,*r*_ is the breast cancer incidence rate in birth cohort *c* at period *p* for county (region) *r*, *A*_*a*_ is the age component for age *a* with *a* = *p* − *c*, *P*_*p*_ is the period component for period *p*, *C*_*c*_ is the cohort component for birth cohort *c*, and *ns*(…) denotes the natural cubic splines functions.

For the age component we specified inner spline knots at age 50, 52, 54, 56, 60, 70 and 80 years (*kn*_*a*_), adding more knots around the age of menopause to increase flexibility for ages at which the effects of some breast cancer risk factors typically may change (1). We used three degrees of freedom for the period component, and for the birth cohort component, both with knots placed at the corresponding quantiles (for *kn*_*p*_at 1997 and 2009, and for *kn*_*c*_at 1929 and 1943). *S*_1_, *S*_2_, *S*_3+_ and *S*_*post*_are coefficients for *scr*_1_, *scr*_2_, *scr*_3+_and *scr*_*post*_, respectively, reflecting initial screening, second screening, continued screening and post screening. For the natural cubic splines applied to the age component for subsequent screening rounds, we specified an inner spline knot at 60 years of age, and boundary knots at ages 53 and 72 years (*kn*_*Aa*∗*scr*3+_). *timeScr*_*post*_is the time since screening cessation, given as mean value over each calendar year, and we specified inner spline knots at 1, 2, 5, 10 and 13 years (*kn*_*scrpost*_) for this variable. *H* is the coefficient for the hormone therapy variable *ht*. Each county was assigned its own level, as given by the *County* variable. The logarithm of the number of person-years under study was used as offset adjusting for variations in person-years.

**Table S.2.**
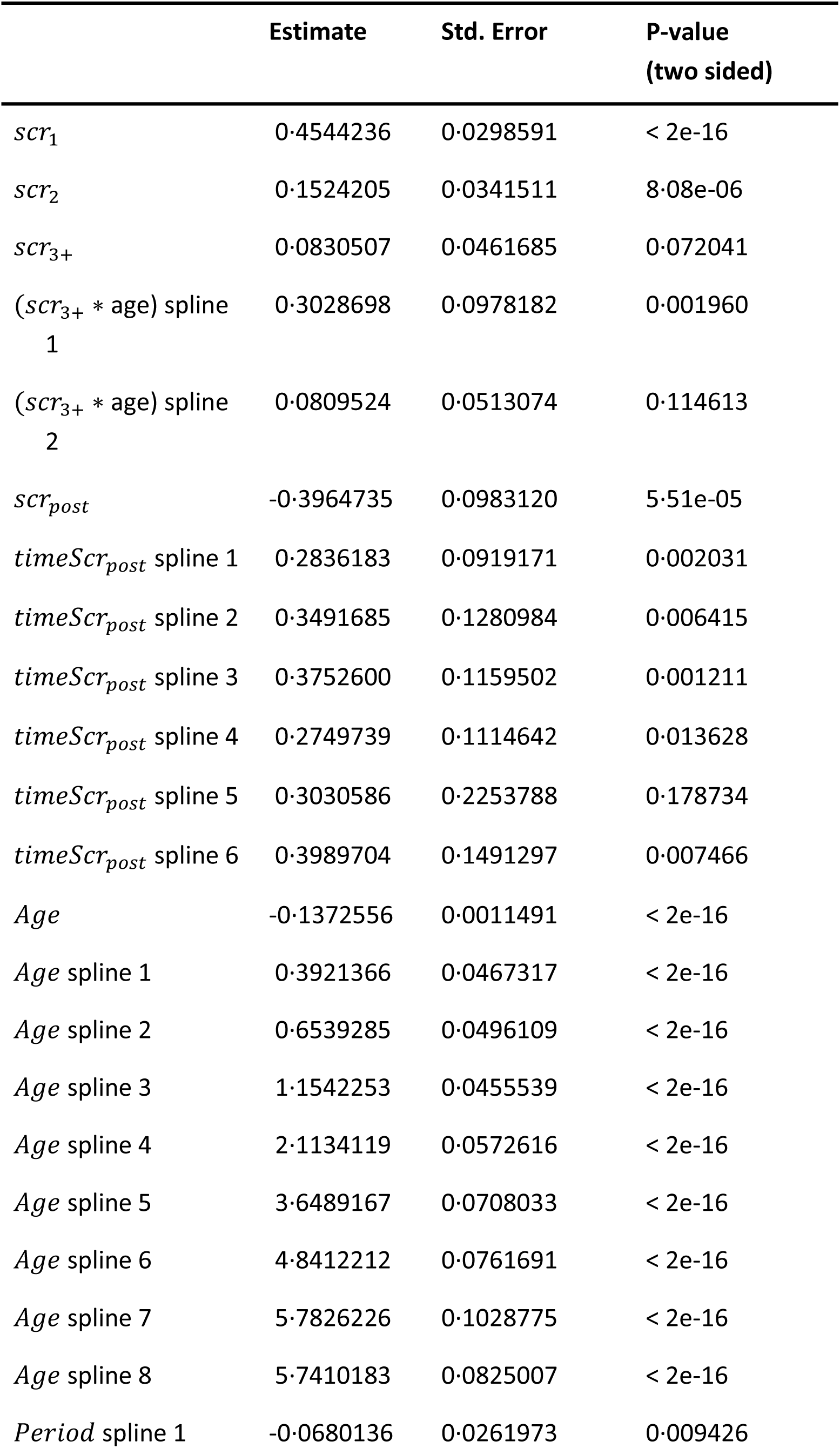
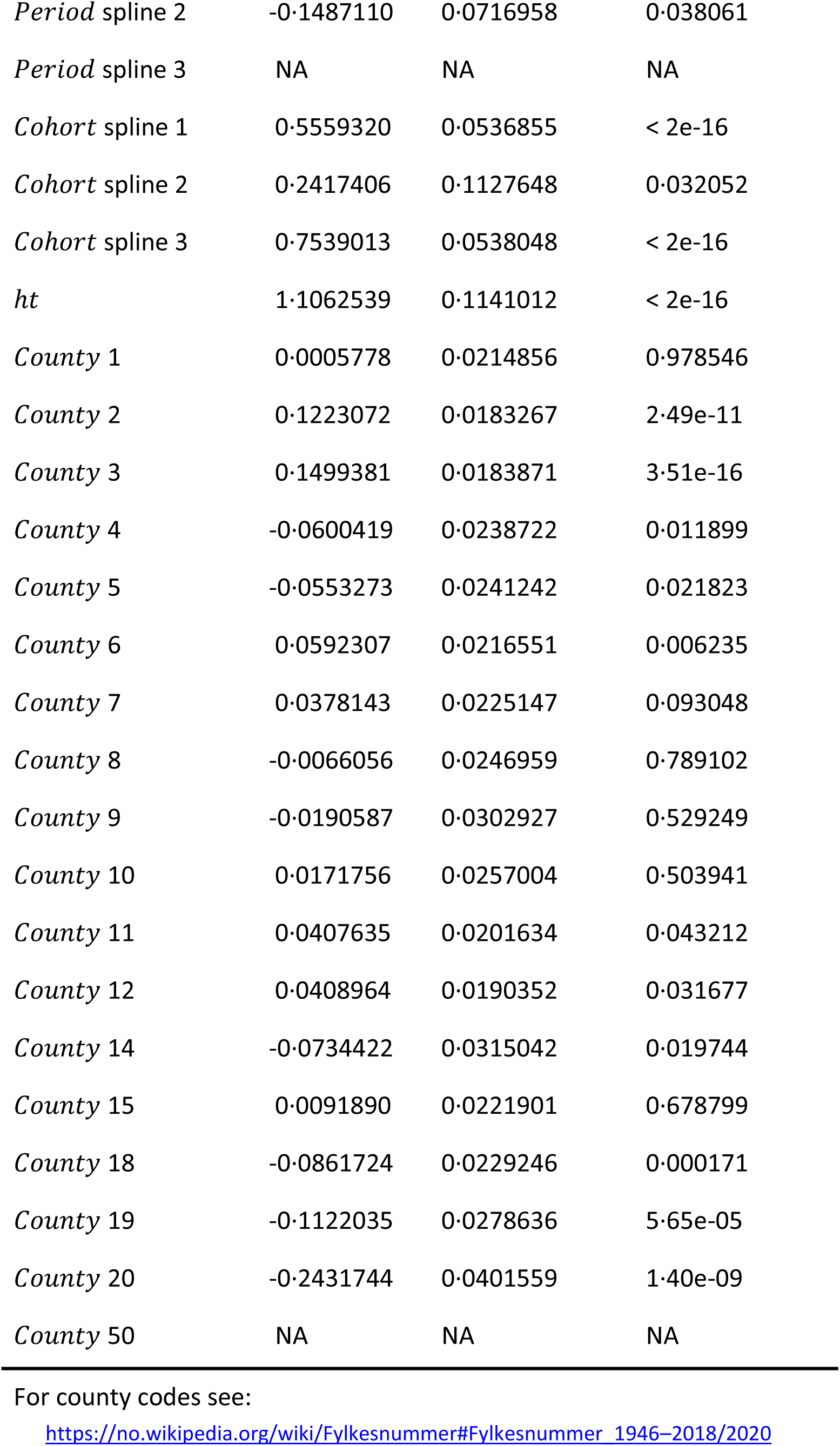
Fitted parameters for the incidence model for age 49-84 years.

|  | Estimate | Std. Error | P-value<br>(two sided) |
| --- | --- | --- | --- |
| $scr_1$ | 0.4544236 | 0.0298591 | < 2e-16 |
| $scr_2$ | 0.1524205 | 0.0341511 | 8.08e-06 |
| $scr_{3+}$ | 0.0830507 | 0.0461685 | 0.072041 |
| ($scr_{3+}$ * age) spline 1 | 0.3028698 | 0.0978182 | 0.001960 |
| ($scr_{3+}$ * age) spline 2 | 0.0809524 | 0.0513074 | 0.114613 |
| $scr_{post}$ | -0.3964735 | 0.0983120 | 5.51e-05 |
| $timeScr_{post}$ spline 1 | 0.2836183 | 0.0919171 | 0.002031 |
| $timeScr_{post}$ spline 2 | 0.3491685 | 0.1280984 | 0.006415 |
| $timeScr_{post}$ spline 3 | 0.3752600 | 0.1159502 | 0.001211 |
| $timeScr_{post}$ spline 4 | 0.2749739 | 0.1114642 | 0.013628 |
| $timeScr_{post}$ spline 5 | 0.3030586 | 0.2253788 | 0.178734 |
| $timeScr_{post}$ spline 6 | 0.3989704 | 0.1491297 | 0.007466 |
| *Age* | -0.1372556 | 0.0011491 | < 2e-16 |
| *Age* spline 1 | 0.3921366 | 0.0467317 | < 2e-16 |
| *Age* spline 2 | 0.6539285 | 0.0496109 | < 2e-16 |
| *Age* spline 3 | 1.1542253 | 0.0455539 | < 2e-16 |
| *Age* spline 4 | 2.1134119 | 0.0572616 | < 2e-16 |
| *Age* spline 5 | 3.6489167 | 0.0708033 | < 2e-16 |
| *Age* spline 6 | 4.8412212 | 0.0761691 | < 2e-16 |
| *Age* spline 7 | 5.7826226 | 0.1028775 | < 2e-16 |
| *Age* spline 8 | 5.7410183 | 0.0825007 | < 2e-16 |
| *Period* spline 1 | -0.0680136 | 0.0261973 | 0.009426 |
| *Period* spline 2 | -0.1487110 | 0.0716958 | 0.038061 |
| *Period* spline 3 | NA | NA | NA |
| *Cohort* spline 1 | 0.5559320 | 0.0536855 | < 2e-16 |
| *Cohort* spline 2 | 0.2417406 | 0.1127648 | 0.032052 |
| *Cohort* spline 3 | 0.7539013 | 0.0538048 | < 2e-16 |
| *ht* | 1.1062539 | 0.1141012 | < 2e-16 |
| *County* 1 | 0.0005778 | 0.0214856 | 0.978546 |
| *County* 2 | 0.1223072 | 0.0183267 | 2.49e-11 |
| *County* 3 | 0.1499381 | 0.0183871 | 3.51e-16 |
| *County* 4 | -0.0600419 | 0.0238722 | 0.011899 |
| *County* 5 | -0.0553273 | 0.0241242 | 0.021823 |
| *County* 6 | 0.0592307 | 0.0216551 | 0.006235 |
| *County* 7 | 0.0378143 | 0.0225147 | 0.093048 |
| *County* 8 | -0.0066056 | 0.0246959 | 0.789102 |
| *County* 9 | -0.0190587 | 0.0302927 | 0.529249 |
| *County* 10 | 0.0171756 | 0.0257004 | 0.503941 |
| *County* 11 | 0.0407635 | 0.0201634 | 0.043212 |
| *County* 12 | 0.0408964 | 0.0190352 | 0.031677 |
| *County* 14 | -0.0734422 | 0.0315042 | 0.019744 |
| *County* 15 | 0.0091890 | 0.0221901 | 0.678799 |
| *County* 18 | -0.0861724 | 0.0229246 | 0.000171 |
| *County* 19 | -0.1122035 | 0.0278636 | 5.65e-05 |
| *County* 20 | -0.2431744 | 0.0401559 | 1.40e-09 |
| *County* 50 | NA | NA | NA |

For county codes see:
https://no.wikipedia.org/wiki/Fylkesnummer#Fylkesnummer_1946–2018/2020

### Estimated parameters for the applied breast carcinoma incidence model

### Pre-menopause breast carcinoma incidence model

For estimating increased lifetime breast carcinoma risk, breast cancer incidence pre 49 years of age is needed. Hence, we fitted a separate model for women up to 48 years of age. No organized screening takes place for women in this age group and our model was given by:

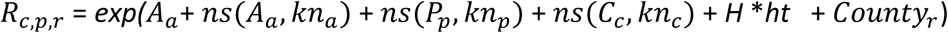

where *R*_*c*,*p*,*r*_ is the breast cancer incidence rate in birth cohort *c* at period *p* for county(region) *r*, *A*_*a*_is the age component for age *a* = *p* − *c*, *P*_*p*_is the period component for period *p*, and *C*_*c*_ is the cohort component for birth cohort *c*. *ns*(…) denotes a natural cubic splines function. For the age component we specified inner spline knots at age 10, 20, 30, 35, 40 and 45 years (*kn*_*a*_). We used three degrees of freedom for the period component, and three for the birth cohort component, both with knots set at the corresponding quantiles defined as (*kn*_*p*_and *kn*_*c*_, respectively). As for the main model, *H* is the coefficient for the hormone therapy variable *ht*. Each county got its own level given by the *County* variable, while the logarithm of the number of person-years under study was used as offset adjusting for variations in person-years.

### Pre-hoc breast carcinoma incidence model

To make sure our modelling choices are not influenced by pre-study assumptions regarding likely over-diagnosis levels, we specified a model pre-hoc before doing any analysis. The pre-hoc model included women from the year they turned 48 years. The reason for this choice was to include all cancers detected in BreastScreen Norway, as some women might be invited to screening before they turn 49 years of age. However, as the data were organized by calendar year and birth cohort, we later realized that no BreastScreen women were defined as 48 years of age in our data. As for model specification, the pre-hoc model was defined as:

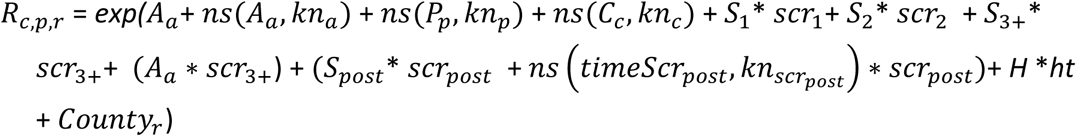

where *R*_*c*,*p*,*r*_ is the breast cancer incidence rate in birth cohort *c* at period *p* for county(region) *r*, *A*_*a*_is the age component for age *a* = *p* − *c*, *P*_*p*_is the period component for period *p*, and *C*_*c*_ is the cohort component for birth cohort *c*, and *ns*(…) denotes a natural cubic splines function. For the age component we specified inner spline knots at age 50, 52, 54, 56, 60, 70 and 80 years (*kn*_*a*_). We used three degrees of freedom for the period and birth cohort components, both with knots set at the corresponding quantiles (defined as *kn*_*p*_and *kn*_*c*_, respectively). *S*_1_, *S*_2_, *S*_3+_ and *S*_*post*_are coefficients for the corresponding *scr*_1_, *scr*_2_, *scr*_3+_and *scr*_*post*_, screening variables. *scr*_1_reflects initial screening, *scr*_2_first subsequent screening, *scr*_3+_continued screening, and *scr*_*post*_previous screening. For previous screening *timeScr*_*post*_is the time since screening ceased, and we specified inner spline knots at 2, 5, 10 and 13 years (*kn*_*scrpost*_) for *timeScr*_*post*_. In addition, *H* is the coefficient for the hormone therapy variable *ht*. Each county got its own level as given by the *County* variable, while the logarithm of the number of person-years under study was used as offset adjusting for variations in person-years.

In addition to changing the age range to 49-84 years in our applied model, we made the following changes to the pre-hoc model: i) a spline knot at 1 year was added for *timeScr*_*post*_ to increase the flexibility of the model, and ii) natural cubic splines were added to continued screening to allow for non-linear effects by age for subsequent screening rounds.

### Calculation of lifetime risk of breast carcinoma

When estimating the lifetime risk of a breast cancer diagnosis with and without screening, the key is to align the applied definitions of being under risk of developing breast cancer across the estimation model and the calculation of estimated lifetime risk. If only women without a previous diagnosis are included in the incidence model, this must correspondingly be adjusted for when cumulating the estimated age-specific risks. Defining *R*_*i*_ as the risk of a new event at age *i*, and *S*_*i*_as the probability of still being under study at age *i*, we might either:

a. Define *R*_*i*_ as the age-specific risk of a breast carcinoma diagnosis among all women alive at age *i* with no previous breast carcinoma diagnosis, and *S*_*i*_as the probability of being alive with no previous breast carcinoma diagnosis by age *i* or
b. Define *R*_*i*_as the age-specific risk of a first breast carcinoma diagnosis among all women alive at age *i*, and *S*_*i*_as the probability of being alive at age *i*

Due to the format of the given Norwegian data, we chose here option (b), where all women who were alive were included in the risk set at a given age.

### County-specific breast cancer incidence rate

**Figure S.2.**
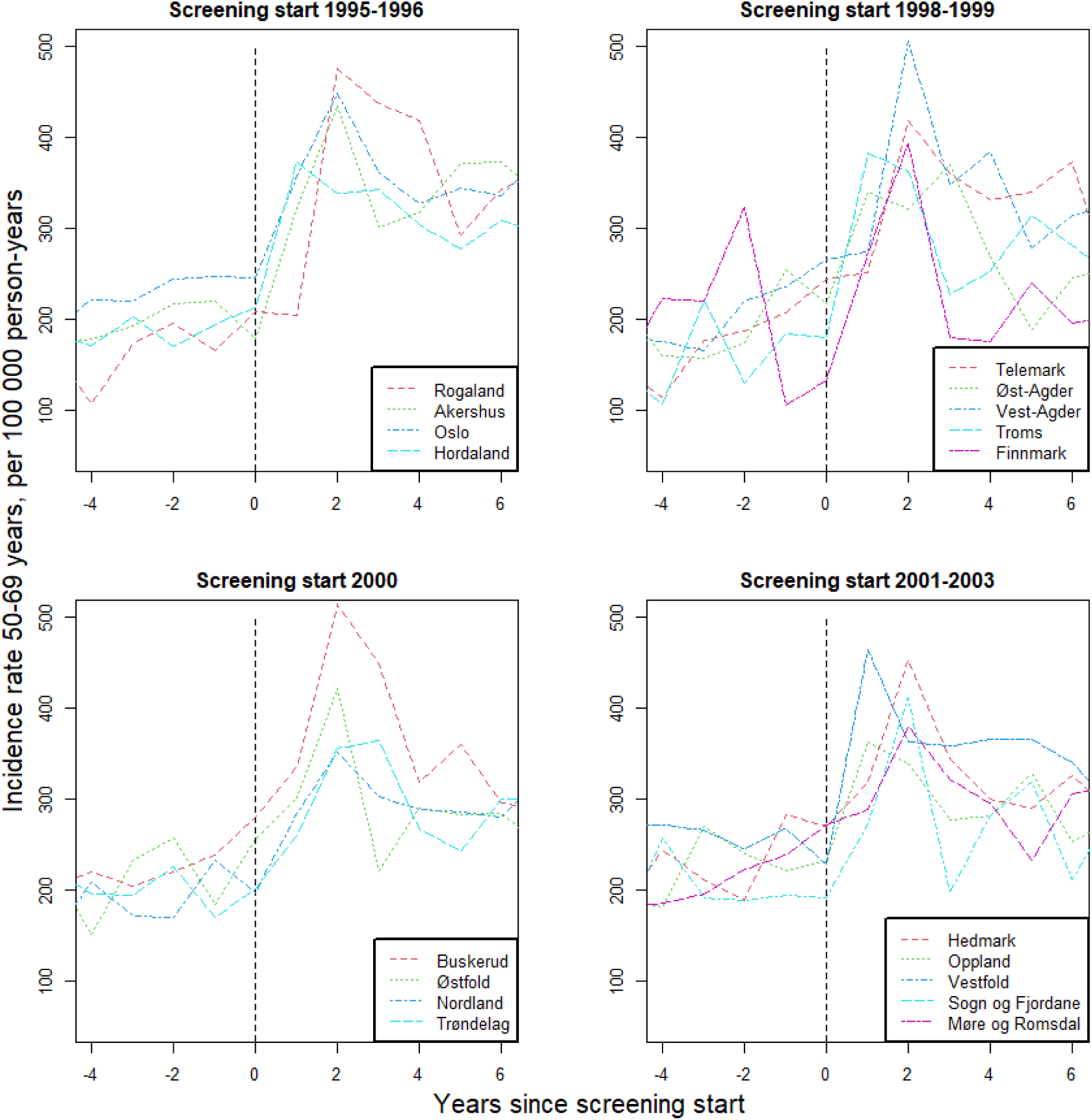
Breast cancer incidence rates for women aged 50-69 years in Norwegian counties. The dotted line indicate the year of county-specific screening initiation.

## References

1. Marmot MG, Altman DG, Cameron DA, et al. The benefits and harms of breast cancer screening: an independent review. Br J Cancer 2013; 108: 2205–2240. 2013/06/08. DOI: 10.1038/bjc.2013.177.

2. Lund E, Nakamura A and Thalabard JC. No overdiagnosis in the Norwegian Breast Cancer Screening Program estimated by combining record linkage and questionnaire information in the Norwegian Women and Cancer study. Eur J Cancer 2018; 89: 102–112. 2017/12/16. DOI: 10.1016/j.ejca.2017.11.003.

3. Ryser MD, Lange J, Inoue LYT, et al. Estimation of Breast Cancer Overdiagnosis in a U.S. Breast Screening Cohort. Ann Intern Med 2022 2022/03/01. DOI: 10.7326/m21-3577.

4. Jørgensen KJ, Zahl PH and Gøtzsche PC. Overdiagnosis in organised mammography screening in Denmark. A comparative study. BMC Womens Health 2009; 9: 36. 2009/12/24. DOI: 10.1186/1472-6874-9-36.

5. Bleyer A and Welch HG. Effect of three decades of screening mammography on breast-cancer incidence. The New England journal of medicine 2012; 367: 1998–2005. DOI: 10.1056/NEJMoa1206809.

6. Zahl PH, Strand BH and Maehlen J. Incidence of breast cancer in Norway and Sweden during introduction of nationwide screening: prospective cohort study. BMJ 2004; 328: 921–924. DOI: 10.1136/bmj.38044.666157.63.

7. Ding L, Poelhekken K, Greuter MJW, et al. Overdiagnosis of invasive breast cancer in population-based breast cancer screening: A short- and long-term perspective. Eur J Cancer 2022; 173: 1–9. 20220712. DOI: 10.1016/j.ejca.2022.06.027.

8. de Gelder R, Heijnsdijk EA, van Ravesteyn NT, et al. Interpreting overdiagnosis estimates in population-based mammography screening. Epidemiol Rev 2011; 33: 111–121. DOI: 10.1093/epirev/mxr009.

9. Njor S, Paci E and Rebolj M. As you like it: How the same data can support manifold views of overdiagnosis in breast cancer. International journal of Cancer 2018; 143: 1287–1294. DOI: 10.1002/ijc.31420.

10. Njor SH, Garne JP and Lynge E. Over-diagnosis estimate from The Independent UK Panel on Breast Cancer Screening is based on unsuitable data. J Med Screen 2013; 20: 104–105. 2013/09/26. DOI: 10.1177/0969141313495190.

11. Larsen IK, Smastuen M, Johannesen TB, et al. Data quality at the Cancer Registry of Norway: an overview of comparability, completeness, validity and timeliness. Eur J Cancer 2009; 45: 1218–1231. 2008/12/19. DOI: 10.1016/j.ejca.2008.10.037.

12. Hofvind S, Tsuruda K, Mangerud G, et al. The Norwegian Breast Cancer Screening Program, 1996-2016: Celebrating 20 years of organised mammographic screening. In: Cancer in Norway 2016 - Cancer incidence, mortality, survival and prevalence in Norway. Oslo: Cancer Registry of Norway, 2017.

13. Hofvind S, Skaane P, Elmore JG, et al. Mammographic performance in a population-based screening program: before, during, and after the transition from screen-film to full-field digital mammography. Radiology 2014; 272: 52–62. 2014/04/03. DOI: 10.1148/radiol.14131502.

14. Weedon-Fekjaer H, Bakken K, Vatten LJ, et al. Understanding recent trends in incidence of invasive breast cancer in Norway: age-period-cohort analysis based on registry data on mammography screening and hormone treatment use. BMJ 2012; 344: e299. 2012/02/01. DOI: 10.1136/bmj.e299.

15. Bakken K, Alsaker E, Eggen AE, et al. Hormone replacement therapy and incidence of hormone-dependent cancers in the Norwegian Women and Cancer study. Int J Cancer 2004; 112: 130–134. 2004/08/12. DOI: 10.1002/ijc.20389.

16. Rossouw JE, Anderson GL, Prentice RL, et al. Risks and benefits of estrogen plus progestin in healthy postmenopausal women: principal results From the Women’s Health Initiative randomized controlled trial. JAMA 2002; 288: 321–333. 2002/07/19. DOI: 10.1001/jama.288.3.321.

17. Norwegian Institute of Public Health. WHO Collaborating Centre for Drug Statistics Methodology. https://www.whocc.no (accessed March 7 2024).

18. Clayton D and Schifflers E. Models for temporal variation in cancer rates. I: Age-period and age-cohort models. Stat Med 1987; 6: 449–467. 1987/06/01. DOI: 10.1002/sim.4780060405.

19. Clayton D and Schifflers E. Models for temporal variation in cancer rates. II: Age-period-cohort models. Stat Med 1987; 6: 469–481. 1987/06/01. DOI: 10.1002/sim.4780060406.

20. Bell A. Age period cohort analysis: a review of what we should and shouldn’t do. Ann Hum Biol 2020; 47: 208–217. DOI: 10.1080/03014460.2019.1707872.

21. Moller B, Weedon-Fekjaer H, Hakulinen T, et al. The influence of mammographic screening on national trends in breast cancer incidence. Eur J Cancer Prev 2005; 14: 117–128.

22. Heggland T, Vatten LJ, Opdahl S, et al. Non-progressive breast carcinomas detected at mammography screening: a population study. Breast cancer research: BCR 2023; 25: 80. 20230704. DOI: 10.1186/s13058-023-01682-9.

23. Heggland T, Vatten LJ, Opdahl S, et al. Non-progressive breast carcinomas detected at mammography screening: a population study [Supplemental material]. Breast Cancer Res 2023; 25: 80. 20230704. DOI: 10.1186/s13058-023-01682-9.

24. Heer E, Harper A, Escandor N, et al. Global burden and trends in premenopausal and postmenopausal breast cancer: a population-based study. Lancet Glob Health 2020; 8: e1027–e1037. 2020/07/28. DOI: 10.1016/s2214-109x(20)30215-1.

25. R Developer Core Team. R: A language and environment for statistical computing. R Foundation for Statistical Computing, Vienna, Austria. 2022.

26. Weedon-Fekjaer H, Romundstad PR and Vatten LJ. Modern mammography screening and breast cancer mortality: population study. BMJ 2014; 348: g3701. 2014/06/22. DOI: 10.1136/bmj.g3701.

27. Duffy SW, Cuzick J, Tabar L, et al. Correcting for Non-Compliance Bias in Case-Control Studies to Evaluate Cancer Screening Programmes Journal of the Royal Statistical Society Series C (Applied Statistics) 2002; 51: 235–243. DOI: 10.1111/1467-9876.00266.

28. Spix C, Berthold F, Hero B, et al. Correction factors for self-selection when evaluating screening programmes. J Med Screen 2016; 23: 44–49. 20150729. DOI: 10.1177/0969141315597959.

29. Weedon-Fekjaer H, Lindqvist BH, Vatten LJ, et al. Estimating mean sojourn time and screening sensitivity using questionnaire data on time since previous screening. Journal of medical screening 2008; 15: 83–90. DOI: 10.1258/jms.2008.007071.

30. Falk RS, Hofvind S, Skaane P, et al. Overdiagnosis among women attending a population-based mammography screening program. Int J Cancer 2013; 133: 705–712. 2013/01/29. DOI: 10.1002/ijc.28052.

31. Hofvind S, Vacek PM, Skelly J, et al. Comparing screening mammography for early breast cancer detection in Vermont and Norway. J Natl Cancer Inst 2008; 100: 1082–1091. 2008/07/31. DOI: 10.1093/jnci/djn224.

32. Azam S, Jacobsen KK, Aro AR, et al. Hormone replacement therapy and mammographic density: a systematic literature review. Breast Cancer Res Treat 2020; 182: 555–579. 20200622. DOI: 10.1007/s10549-020-05744-w.

33. Heinig M, Schwarz S and Haug U. Self-selection for mammography screening according to use of hormone replacement therapy: A systematic literature review. Cancer Epidemiol 2021; 71: 101812. 20210216. DOI: 10.1016/j.canep.2020.101812.

34. Farber R, Houssami N, Wortley S, et al. Impact of Full-Field Digital Mammography Versus Film-Screen Mammography in Population Screening: A Meta-Analysis. Journal of the National Cancer Institute 2021; 113: 16–26. DOI: 10.1093/jnci/djaa080.

35. Lynge E, Braaten T, Njor SH, et al. Mammography activity in Norway 1983 to 2008. Acta Oncol 2011; 50: 1062–1067. Research Support, Non-U.S. Gov’t 2011/08/13. DOI: 10.3109/0284186X.2011.599339.

36. Kalager M, Adami HO, Bretthauer M, et al. Overdiagnosis of invasive breast cancer due to mammography screening: results from the Norwegian screening program. Annals of internal medicine 2012; 156: 491–499. DOI: 10.1059/0003-4819-156-7-201204030-00005.

37. Ripping TM, Verbeek AL, Fracheboud J, et al. Overdiagnosis by mammographic screening for breast cancer studied in birth cohorts in The Netherlands. Int J Cancer 2015; 137: 921–929. 2015/01/24. DOI: 10.1002/ijc.29452.

38. Njor SH, Olsen AH, Blichert-Toft M, et al. Overdiagnosis in screening mammography in Denmark: population based cohort study. BMJ 2013; 346. DOI: 10.1136/bmj.f1064.

39. Blyuss O, Dibden A, Massat NJ, et al. A case-control study to evaluate the impact of the breast screening programme on breast cancer incidence in England. Cancer medicine 2023; 12: 1878–1887. 20220718. DOI: 10.1002/cam4.5004.

40. van Luijt PA, Heijnsdijk EA, van Ravesteyn NT, et al. Breast cancer incidence trends in Norway and estimates of overdiagnosis. J Med Screen 2017; 24: 83–91. 2016/10/19. DOI: 10.1177/0969141316668379.

41. de Koning HJ, Draisma G, Fracheboud J, et al. Overdiagnosis and overtreatment of breast cancer: microsimulation modelling estimates based on observed screen and clinical data. Breast Cancer Res 2006; 8: 202. 2006/03/10. DOI: 10.1186/bcr1369.

42. Michalopoulos D and Duffy SW. Estimation of overdiagnosis using short-term trends and lead time estimates uncontaminated by overdiagnosed cases: Results from the Norwegian Breast Screening Programme. J Med Screen 2016; 23: 192–202. 2016/03/05. DOI: 10.1177/0969141315623980.

## References

1. Heer E, Harper A, Escandor N, et al. Global burden and trends in premenopausal and postmenopausal breast cancer: a population-based study. Lancet Glob Health 2020;8(8):e1027–e37. doi: 10.1016/s2214-109x(20)30215-1 [published Online First: 2020/07/28]

